# Measurement and interpretation of the Harare HIV combination prevention cascade in priority populations: A population survey of adolescent girls and young women and young men in Zimbabwe

**DOI:** 10.1101/2025.01.08.25320195

**Authors:** Louisa Moorhouse, Jeffrey W. Imai-Eaton, Tawanda Dadirai, Rufurwokuda Maswera, Tafadzwa Museka, Phyllis Mandizvidza, Freedom Dzamatira, Blessing Tsenesa, Timothy B Hallett, Constance Nyamukapa, Simon Gregson

## Abstract

**Introduction:** HIV-negative adolescent girls and young women (AGYW), and male partners, have disproportionately high HIV incidence in many African countries. We used a new HIV Prevention Cascade (HPC) approach to quantify levels of, and barriers to, prevention method use to guide interventions to increase effective uptake of primary HIV prevention.

**Methods:** Data from the Manicaland HPC pilot study (2018-19; N=9803) in Zimbabwe were used to measure levels of sexual risk behaviour and construct HPCs for male condom, PrEP (females), VMMC (males) and combination prevention use by HIV-negative sexually-active AGYW (15-24-years) and male partners (15-29-years).

**Results:** 19% of AGYW (N=1140) and 37% of young men (N=955) who had started sex reported one or more HIV risk behaviour and met the definition of the priority populations for HIV prevention. Of these, 63% of females and 87% of males were motivated to use an HIV prevention method, 28% and 63% had access to a method, and 16% and 53% used a method. Male condoms were the most commonly used prevention method, accounting for 97% of use in females and 55% in males. Barriers to motivation, access and capacity to use were reported for all priority populations and methods. Some barriers were common across HPCs (e.g. lack of risk perception, social unacceptability, and lack of acceptable provision); others were specific to particular prevention methods or priority populations (e.g. lack of availability).

**Conclusion:** HIV risk behaviours were commonly reported, but use of prevention methods is low in young people in Manicaland. Population survey measurements of HPCs revealed large gaps in all steps in the cascade (lack of motivation, lack of access and lack of capacity to use prevention) but also provided information on the reasons for these gaps that can aid in designing interventions that reduce new infections.

## Introduction

HIV incidence has declined in eastern and southern Africa due to protective changes in sexual behaviours^1,2^ and widespread availability of antiretroviral therapy (ART) reducing HIV infectiousness^3,4^. However, reductions in incidence have been slower than targeted and the UNAIDS milestone of reducing global new infections to fewer than 500,000 per year by 2020 was missed^5^ ^6,7^. Incidence declines have varied between population sub-groups across eastern and southern Africa^8^ ^9^. This heterogeneity has been attributed to a combination of variation in the provision of HIV prevention programmes and uptake of preventive behaviour^10^, emphasising the need to improve understanding of why people are not adopting HIV prevention and for novel approaches to improve the use and impact of prevention methods. HIV-negative adolescent girls and young women (AGYW), in particular, experience high HIV incidence in the region and therefore have been identified as a priority population for targeting prevention ^11,12^.

Despite availability of efficacious HIV prevention methods, levels of use vary considerably by type of sexual relationship and between populations at risk of acquiring HIV. When used correctly, condoms reduce HIV transmission by 90-95%^13,14^. Voluntary medical male circumcision (VMMC) reduces HIV acquisition in men by 60%^15^. Oral pre-exposure prophylaxis (PrEP) reduces acquisition risk by up to 90%, with good adherence^16^. However, UNAIDS estimate a gap of 3 billion condoms a year in eastern and southern Africa^6^. While VMMC expanded rapidly in the second half of the 2010s, the number of VMMC procedures reduced by half from 2019 to 2020 with service disruptions following the COVID-19 pandemic^17^. PrEP use is a key tool in the 2025 UNAIDS roadmap for HIV prevention and gaps remain in the provision and demand for PrEP^17^. PrEP targets aim to make PrEP available to all people at elevated risk of HIV infection – estimated to be 10 million people^17^.

Understanding multilevel components contributing to insufficient use of prevention methods, including demand, supply and structural barriers, is crucial to reaching global targets for HIV incidence reduction. The Harare HIV Combination Prevention Cascade (HIV-CPC) has been proposed as a generic framework to be applied to multiple populations and prevention methods^18^, and has been developed through a series of consultations and literature review^19^. It focuses on identification of a priority population in need of HIV prevention methods followed by three core steps, represented as sequential bars in a cascade: motivation to use, access to, and effective use of the prevention method. Large gaps between successive steps in the cascade can be interrogated according to the frequencies of potential barriers that underpin most common reasons for these gaps. The sub-bars hypothesised in the HPC^18^, identified from literature review, social-cognitive models and behavioural and epidemiological theories^20^, represent these barriers and can suggest targets for interventions most likely to be effective at increasing use of the prevention method.

In this study, we carried out a pilot study in east Zimbabwe to measure and interpret the Harare HIV-CPC using data from a general population survey. We previously established that this is feasible and provides valid data on the gaps and barriers to use of prevention methods^21^. In this study, we demonstrate the utility of collecting and interpreting the Harare HIV-CPC in a population survey using the examples of priority populations of AGYW and their potential male partners. In so doing, we provide valuable new insights into the types of interventions needed to increase use of prevention methods and to reduce new infections in these vulnerable groups.

## Methods

### Study setting and data source

Data were from the Manicaland HPC Study, conducted across eight study sites in east Zimbabwe (July 2018-December 2019) which represent urban, peri-urban, farming estates and subsistence farming areas. A household census was conducted and household members, aged ≥15 years, were invited to participate in individual interviews. Data on socio-demographic characteristics, HIV knowledge, risk and prevention method use were collected using a questionnaire designed specifically to populate HPCs. Sensitive questions were asked using a secret voting method^22^.

All participants completing the individual questionnaire received HIV counselling and testing (HTC) and were requested to provide a dried blood spot (DBS) sample for laboratory testing. HTC was conducted using the Zimbabwe Ministry of Health’s rapid testing algorithm and guidelines, based on the 2015 WHO recommendations^23^. Where HTC was accepted, the HTC result was used as the final HIV status result. Where HTC was not completed but consent and DBS was provided for laboratory testing, the same testing algorithm was completed at the Biomedical Research and Training Institute laboratory using the DBS.

### Data analysis

Data were restricted to HIV-negative females aged 15-24 years and males aged 15-29 years. Proportions and 95% confidence intervals were calculated for socio-demographic characteristics. Socioeconomic status was calculated as a wealth score taking into account reported sellable and non-sellable household assets split into quintiles^24^. Descriptive statistics for participants’ sexual risk behaviours were calculated for survey participants who self-reported having ever had sex.

Harare HPCs^18,19^ were populated separately for males and females. The female priority population for the cascades was defined as HIV-negative women aged 15-24 years who self-reported at least one risk behaviour in the last 12 months. The male partner priority population was defined as HIV-negative men aged 15-29 years self-reporting ≥1 sexual risk behaviour in the last 12 months. This age-range for potential male partners was based on the age-range of partners most commonly reported by young women in previous surveys in the study areas. Risk behaviours (identified through previous analyses of Manicaland cohort data^25^) were having multiple partners in the last 12 months; concurrent partners at the time of interview; recent transactional sex in the last month with any of the last three partners; and reporting ≥1 non-regular partner in the last 12 months.

To create the main bars in the HPCs for individual HIV prevention methods, proportions and 95% CIs of the priority populations reporting motivation, access and effective use were calculated separately for male condoms, female condoms, PrEP and VMMC. If an individual reported currently using male condoms, female condoms, or PrEP as a method of preventing HIV, they were defined as effectively using the respective prevention method. Effective use of VMMC was defined as having taken up full medical male circumcision. Exact definitions and full methods for populating the cascade have been validated and published separately^21^. Individuals who reported effectively using a particular HIV prevention method were assumed to be motivated and have access to that method.

Full extended HPCs reflecting explanatory factor sub-bars were created for male condoms, female condoms, PrEP, and VMMC (men only). The frequencies of each of the explanatory factors among individuals who reported gaps in the main HPCs for each prevention method were measured and shown as sub-bars in the expanded cascade diagrams. Logistic regression was used to assess associations between sociodemographic characteristics and prevention method use, and differences between the HPCs.

HIV combination prevention cascades were created to assess motivation, access, and use of at least one prevention method for males (VMMC, male condoms, female condoms) and for females (PrEP, male condoms, female condoms). Stacked bar combination HPCs were created: firstly, with levels of motivation, access, and effective use as proportions of the priority population; and then also broken down into each prevention method as proportions of each bar (motivation, access, effective use). Proportions of the priority population reporting motivation, access, and effective use of at least one prevention method (male condoms, female condoms, PrEP, VMMC) were calculated. Ethics approval for the study was granted by the Imperial College Research Ethics Committee and the Medical Research Council of Zimbabwe. Analyses were carried out using Stata MP 17. Tableau was used for data visualisations.

## Results

### Study population and HIV prevalence

Seventy-eight percent (9803/12647) of all eligible individuals completed the individual questionnaire. An HIV result was established for 95% (9339/9803) of individuals completing the individual questionnaire. Forty-six percent (4286/9339) were adolescent and young people (AYP) – 15-24-year-old women or 15-29-year-old men. HIV prevalence among AYP was estimated at 2.76% (95% CI:2.14-3.55) in adolescent boys and young men (ABYM) and 3.12% (95% CI:2.46-3.94) in adolescent girls and young women (AGYW).

### Sociodemographic characteristics

Approximately half of AYP participants were 15-19-years-olds for both men (48%) and women (53%) (Table 1). More young women than young men resided in urban sites (23% vs. 17%) and were currently married (43% vs. 25%). Around 90% of both young men and young women reported secondary or higher education. More than 50% of individuals lived in households in the poorest or second poorest socioeconomic quintile.

**Table 1.** Sociodemographic characteristics and self-reported sexual risk behaviours of HIV-negative adolescent and young people.

|  |  | Female | Male |
| --- | --- | --- | --- |
|  |  | 15-24 years | 15-29 years |
|  |  | N = 2081 | N = 2079 |
|  |  | % (95% CI) | % (95% CI) |
| **Sociodemographic characteristics** |  |  |  |
| 5-year age group |  |  |  |
|  | 15-19 years | 53.4 (51.3-55.6) | 47.7 (45.3-49.6) |
|  | 20-24 years | 45.6 (44.4-48.7) | 31.6 (29.6-33.6) |
|  | 25-29 years | - | 21.0 (19.3-22.8) |
| Site type |  |  |  |
|  | Urban | 23.1 (21.4-25.0) | 17.4 (15.8-19.1) |
|  | Peri-urban | 26.4 (24.6-28.4) | 22.0 (20.3-23.8) |
|  | Estates | 21.6 (19.9-23.5) | 23.0 (28.0-32.0) |
|  | Rural | 28.8 (26.9-30.8) | 30.7 (28.7-32.7) |
| Education |  |  |  |
|  | None/primary | 11.3 (10.1-12.8) | 8.9 (7.7-10.2) |
|  | Secondary/higher | 88.7 (87.2-89.0) | 91.2 (89.9-92.3) |
| Marital status |  |  |  |
|  | Never married | 51.6 (19.5-53.8) | 73.7 (71.8-75.6) |
|  | Currently married | 42.9 (40.8-45.0) | 24.8 (23.0-26.7) |
|  | Divorced/separated | 5.3 (4.4-6.3) | 1.4 (1.0-2.0) |
|  | Widowed | 0.2 (0.1-0.6) | 0.1 (0.0-0.3) |
| Socioeconomic status |  |  |  |
|  | Poorest | 8.6 (7.4-9.9) | 8.8 (7.7-10.1) |
|  | 2nd poorest | 39.5 (37.4-41.6) | 47.7 (45.6-49.9) |
|  | 3rd poorest | 24.8 (23.0-26.8) | 21.7 (20.0-23.5) |
|  | 4th poorest | 25.6 (23.8-27.5) | 20.4 (18.7-22.1) |
|  | Least poor | 1.5 (1.1-2.2) | 1.4 (1.0-2.1) |
| **Sexual Risk Behaviours** |  |  |  |
|  | Number reporting sexual debut<sup>¶</sup> | 1140 | 955 |
|  | Had sexual debut | 54.8 (52.6-56.9) | 45.9 (43.8-48.1) |
|  | Age at first sex <18yrs* | 46.1 (43.3-49.1) | 33.0 (30.1-36.0) |
|  | Had multiple partners in last 12 months* | 4.6 (3.5-5.9) | 21.5 (19.0-24.2) |
|  | Concurrent partners* | 1.1 (0.7-2.0) | 6.7 (5.3-8.5) |
|  | 1 or more non-regular partners in last 12 months* | 13.3 (11.4-15.3) | 31.8 (28.9-34.9) |
|  | Ever engaged in transactional sex* | 8.7 (7.2-10.5) | 14.7 (12.6-17.1) |
|  | Recent transactional sex in last month with any of last 3 partners* | 8.0 (6.5-9.7) | 7.2 (5.7-9.1) |
|  | Median age of last sexual partner<sup>¶</sup> | 26.5 | 20 |
|  | 1 or more of above risk behaviours<sup>¥</sup>* | 18.5 (16.4-20.9) | 37.1 (34.1-40.2) |
|  | 2 or more of above risk behaviours<sup>¥</sup>* | 4.2 (3.2-5.5) | 10.6 (8.8-12.7) |

<sup>¶</sup> reported as actual number not %
\* restricted to those who have had sexual debut
<sup>¥</sup> combining recent transactional sex, non-regular partners, multiple partners and concurrent partners.

### HIV risk behaviours

Fifty-five percent of AGYW (15-24 years) and 46% of ABYM (15-29 years) had started sex (Table 1). Amongst those who had started sex, 22% of ABYM reported multiple partners in the last 12 months compared to 5% for AGYW, and 32% of ABYM reported ≥1 non-regular partners in the last 12 months compared to 13% of AGYW. Few AGYW (1%) reported concurrent partnerships compared to ABYM (6%). Recent transactional sex was similar in AGYW and ABYM (8% vs. 7%). The median age of the last partner reported by females was 26.5 years compared to 20.0 years reported by males. A markedly higher proportion of ABYM reported ≥1 risk behaviour compared to AGYW (37% vs.19%). A total of 37% (n=354) ABYM and 19% (n=211) AGYW met the definition for the prevention priority populations for HPCs.

### Associations of main bars with socio-demographic characteristics

Table 2 shows bivariate associations of socio-demographic characteristics with prevention use among the priority population at risk of HIV infection. Condom use was not significantly associated with age. Men aged 25-29 years had significantly lower odds of having VMMC compared to men aged 15-19 years (OR=0.39, 95% CI:0.18-0.83). Men who had completed secondary or higher education had 2.5 times the odds (95% CI:1.18-5.30) of using male condoms compared to those with no or primary education only. However, VMMC did not vary by education. Male condom use among AGYW did not vary by education level. Being currently married was associated with lower odds of male condom use compared to never being married in both men (OR=0.48, 95% CI:0.31-0.73) and women (OR=0.06, 95% CI:0.02-0.19). Prevention method use did not vary significantly by socioeconomic status.

**Table 2.** Unadjusted associations between sociodemographic characteristics and prevention method use in women and men in the priority population for HIV prevention cascades.

|  |  | Male condoms |  |  |  | VMMC |  |
| --- | --- | --- | --- | --- | --- | --- | --- |
|  |  | Male (N=354) |  | Female (N=211) |  | Male (N=354) |  |
|  |  | Odds ratio (95% CI) | p-value | Odds ratio (95% CI) | p-value | Odds ratio (95% CI) | p-value |
| 5-year age group |  |  |  |  |  |  |  |
|  | 15-19 years | 1.00 |  | 1.00 |  | 1.00 |  |
|  | 20-24 years | 0.86 (0.45-1.64) | 0.637 | 0.96 (0.51-1.82) | 0.912 | 0.63 (0.32-1.23) | 0.177 |
|  | 25-29 years | 1.04 (0.52-2.08) | 0.901 | - | - | 0.39 (0.18-0.83) | 0.014 |
| Site type |  |  |  |  |  |  |  |
|  | Urban | 1.00 |  | 1.00 |  | 1.00 |  |
|  | Peri-urban | 0.92 (0.47-1.81) | 0.808 | 0.82 (0.34-2.01) | 0.670 | 1.29 (0.58-2.86) | 0.525 |
|  | Estates | 0.79 (0.43-1.46) | 0.458 | 1.12 (0.48-2.62) | 0.798 | 1.33 (0.65-2.74) | 0.437 |
|  | Rural | 0.79 (0.41-1.54) | 0.651 | 0.52 (0.22-1.24) | 0.139 | 1.31 (0.58-2.95) | 0.519 |
| Education |  |  |  |  |  |  |  |
|  | None/primary | 1.00 |  | 1.00 |  | 1.00 |  |
|  | Secondary/higher | 2.50 (1.18-5.30) | 0.016 | 0.49 (0.22-1.10) | 0.085 | 1.63 (0.60-4.38) | 0.335 |
| Marital status |  |  |  |  |  |  |  |
|  | Never married | 1.00 |  | 1.00 |  | 1.00 |  |
|  | Currently married | 0.48 (0.31-0.73) | 0.002 | 0.06 (0.02-0.19) | <0.001 | 0.59 (0.34-1.02) | 0.059 |
|  | Divorced/separated | 1.76 (0.56-5.49) | 0.334 | 1.92 (0.87-4.24) | 0.107 | 0.51 (0.14-1.80) | 0.293 |
|  | Widowed | 1.00 | - | 1.37 (0.08-22.68) | 0.825 | 1.00 |  |
| Socioeconomic status |  |  |  |  |  |  |  |
|  | Poorest | 1.00 |  | 1.00 |  | 1.00 |  |
|  | 2nd poorest | 0.67 (0.28-1.61) | 0.369 | 0.65 (0.23-1.81) | 0.405 | 0.96 (0.38-2.43) | 0.930 |
|  | 3rd poorest | 0.62 (0.24-1.59) | 0.321 | 0.83 (0.28-2.46) | 0.730 | 0.58 (0.21-1.66) | 0.312 |
|  | 4th poorest | 1.23 (0.45-3.34) | 0.687 | 0.87 (0.28-2.65) | 0.802 | 0.69 (0.24-2.00) | 0.495 |
|  | Least poor | 0.30 (0.04-2.13) | 0.227 | 1.00 |  | 0.68 (0.06-7.16) | 0.747 |

### HIV prevention cascades for women

Among HIV-negative AGYW reporting ≥1 risk behaviour, 10% (22/211) reported being motivated to use PrEP (Figure 1A). Lack of knowledge of PrEP (97%) was the largest barrier in women not reporting motivation. A high percentage (95%) of women not motivated also did not perceive a future risk of HIV infection. Of the AGYW who were motivated, 91% (20/22) could not access PrEP with 75% of these reporting lack of availability. No women in the priority population reported PrEP use.

**Figure 1.**
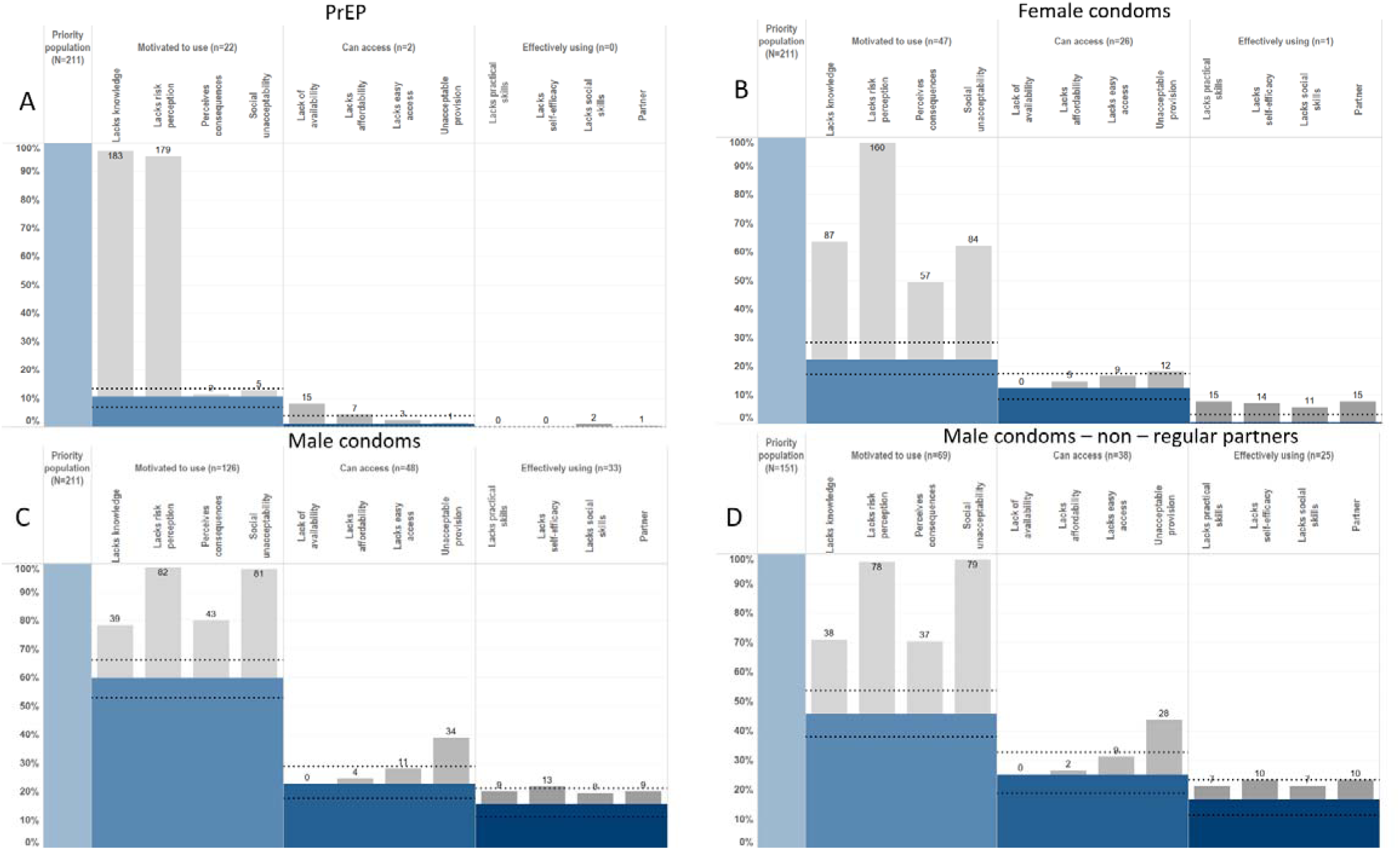
HIV Prevention Cascades for PrEP, Female condom and Male condom use in Young Women. *Blue bars represent proportions reporting motivation, access, and use of a prevention method. Grey bars represent those reporting a barrier to each main bar respectively. Black dashed lines indicate 95% confidence intervals of the proportions motivated, with access and using a prevention method*.

Motivation among AGYW to use female condoms was low (Figure 1B). Only 22% (47/211) reported motivation and <1% reporting using female condoms as an HIV prevention method. Lack of risk perception followed by lack of knowledge and social unacceptability were the biggest barriers in the cascade.

For male condoms, 60% of women were motivated, 23% had access, and 16% were using the method (Figure 1C). Lack of risk perception (96%) was the most common barrier to motivation; however, social unacceptability was also commonly reported (95%). Sixty-two percent of motivated AGYW reported lacking access to male condoms, and 44% of this reported lacking acceptable provision (embarrassment or lack of privacy/confidentiality). Thirty-one percent of women who were motivated and had access to male condoms were not effectively using them. Lack of self-efficacy (including lacking capacity to use condoms due to family or peer disapproval) was the biggest barrier to effective use (87% of those with motivation and access but not using condoms). 60% of those with motivation and access but not effective use reported lacking partner acceptance to use male condoms.

### HIV prevention cascades for men

Among the priority population of HIV-negative ABYM reporting ≥1 risk behaviour, 58% were motivated to use VMMC, 36% could access it, and 23% were fully circumcised (Figure 2A). Fifty-four percent of unmotivated men lacked knowledge of VMMC as an HIV prevention method, 80% did not perceive HIV risk, and 68% perceived negative consequences (painful procedure, irreversible procedure, loss of sexual pleasure). Thirty-nine percent of men motivated to use VMMC could not access it, and 78% of these men reported affordability as a barrier. Of the men who were motivated and had access, 35% were not circumcised and their largest barrier was lack of partner acceptance (36%).

**Figure 2.**
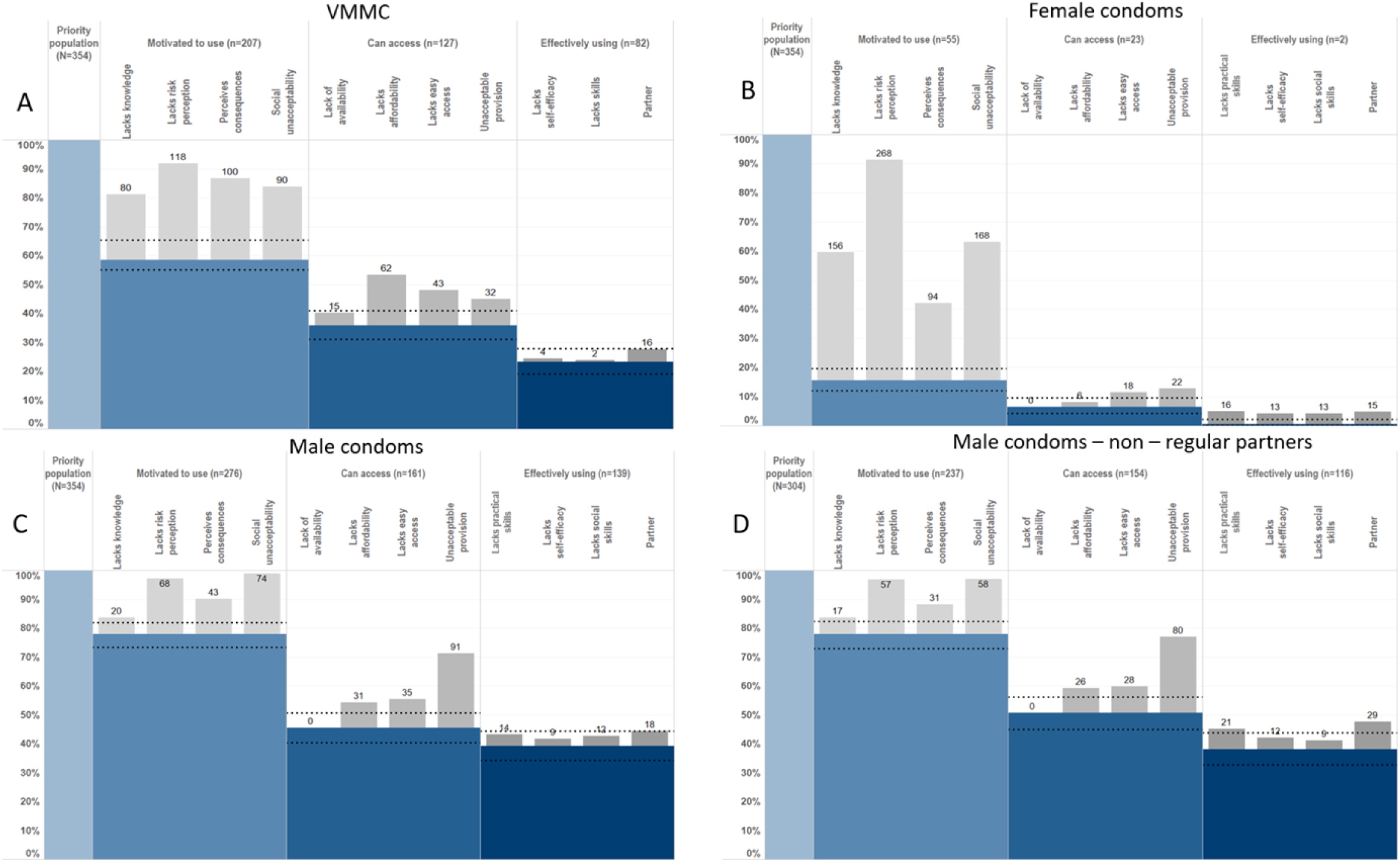
HIV Prevention Cascades for VMMC, Female condom and Male condom use in Young Men*. Blue bars represent proportions reporting motivation, access, and use of a prevention method. Grey bars represent those reporting a barrier to each main bar respectively. Black dashed lines indicate 95% confidence intervals of the proportions motivated, with access and using a prevention method*.

Seventy-eight percent of men were motivated, 45% had access to, and 39% were using male condoms (Figure 2C). Ninety-two percent of men lacking motivation lacked perception of future risk of HIV infection, 55% perceived negative consequences of condom use (reporting reduced sexual pleasure), and 95% reported lack of social acceptability as a barrier to motivation. The drop between motivation and access was the largest drop in this cascade, with 42% of motivated individuals reporting lack of access. Seventy-nine percent of the men who lacked access reported unacceptable provision of male condoms and 27% reported cost as a barrier. Fourteen percent of men who were motivated to use and had access to male condoms were not using them effectively with 82% of these reporting lack of partner acceptance. Lack of self-efficacy to use male condoms was a larger barrier in females than in the equivalent male group: 87% of females vs. 30% of males (OR=9.39; 95% CI:1.69-52.13). Reported use of male condoms was higher in men than in women (OR=1.15; 95% CI:0.59-2.28) and was higher than for PrEP or VMMC, although no statistically significant differences were found. Use of female condoms was low: 16% of men reported motivation to use them and <1% reported use (Figure 2B).

### Combination prevention method use

Overall, 63% of females were motivated, 28% had access to, and 16% were using ≥1 HIV prevention method (Figure 3A. Use of male condoms accounted for 97% of prevention method use (Figure 3B). Of males in the priority population, 87% were motivated to use, 63% had access to, and 53% were using ≥1 method (Figure 3C). Use of male condoms accounted for 55% of prevention method use, followed by VMMC alone (25%) (Figure 3D).

**Figure 3.**
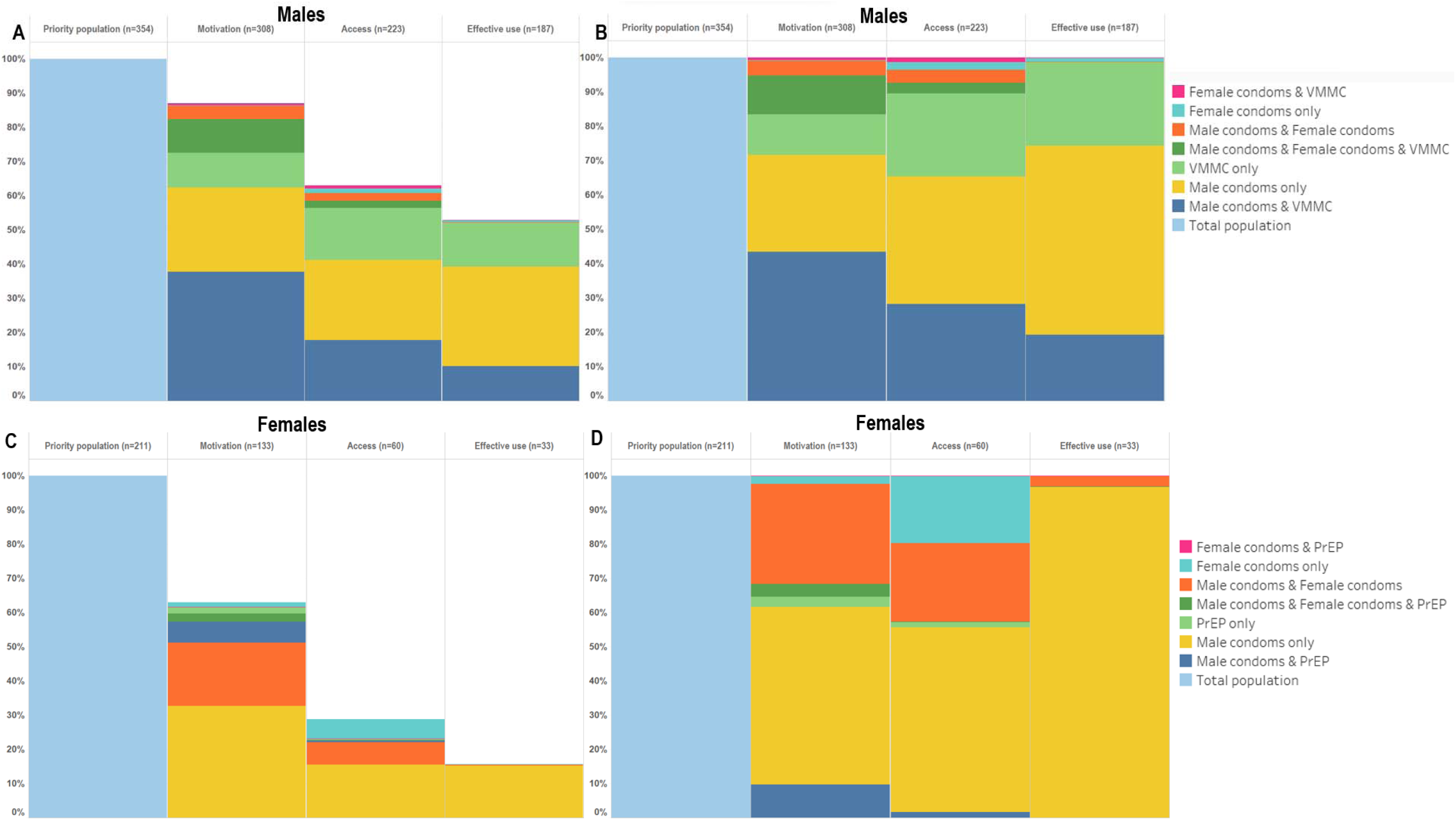
HIV Prevention Cascades showing overall male and female prevention preferences as a proportion of the overall priority population (A&C) and as a proportion of each bar of the cascade (B&D)

## Discussion

This analysis presents the first instance of the Harare HIV-CPC framework being fully populated with general population survey data. Using the Harare HIV-CPC framework, we measured combination and individual primary prevention method use among young people at risk of acquiring HIV, and quantified their particular barriers to individual prevention method use. Barriers were present at all steps of the cascade; indicating that multilevel determinants of prevention method use will need to be targeted by interventions to reduce HIV incidence in young people.

Reported HIV combination prevention cascades were presented in this analysis – one of the first instances of HPCs being applied this way. Levels of motivation, access to and effective use of any prevention method were markedly lower in females than in males with only 16% of females reporting using any prevention method compared to 53% of males. Male condoms remain the most popular (i.e., that people report motivation for), accessible, and widely used primary prevention method, accounting for almost all prevention method use reported. The gap between motivation and access is the largest gap in the combination cascade. Motivation to use ≥1 prevention method is particularly high in young men: 87% of ABYM in the priority population reported wanting to use at least one prevention method. The UNAIDS 2025 Roadmap includes goals of linking at least 90% of people at heightened risk of HIV infection to services, prioritising HIV prevention packages and ensuring they are used by 95% of those at risk of HIV infection^17^. We found that 37% of young men and 19% of young women who have started sex reported ≥1 HIV risk behaviour in the last 12 months, despite declines in risk behaviours observed in earlier studies in Manicaland^2^ although trends in eastern and southern Africa vary^26^. The proportions of the priority populations reporting motivation and access to ≥1 prevention method in this study fall below these targets. Levels of use of prevention methods also remain below targets set out in Zimbabwe’s National HIV and AIDS Strategic Plan^27^, even when assessing use of prevention methods in combination.

Motivation, access and effective use were consistently lower in females than in males for all prevention methods - a key issue given the excess HIV incidence observed in AGYW^11^. This contradicts other reports of generally lower engagement and retention in HIV treatment, testing and prevention services among men in eastern and southern Africa^28^. The reliance on self-reported data may mean estimates of prevention method use were higher due to social desirability biases influencing reporting. Inclusion of VMMC – a one-off procedure - within the male measure of combination prevention may increase the relative estimate of prevention use in men compared to women. The recent availability within PrEP in the study area – and observed lack of knowledge about PrEP – means that reported PrEP use among young women is lower than in other study areas with wider PrEP availability. The Harare HIV-CPC framework provides insight into individual motivation to use prevention methods. The notable number of men lost from the HIV-CPC between the motivation and effective use bars in both young men and women suggests that even where there is demand for primary HIV prevention, other barriers prevent motivated individuals from actually it.

Motivation to use, access to and use of female condoms were low compared to male condoms but comparable to other estimates of female condom use within the region which range from 3% to 38%^29^. Motivation to use, access to and effective use of PrEP were particularly low. Lack of knowledge of PrEP was the biggest barrier to motivation to use, which, together with the reports of poor availability and affordability, reflects the recent introduction of PrEP in Manicaland at the time. Qualitative research carried out in the same population also identified an overall lack of awareness of where to access PrEP^30^. Concerns about disclosure of PrEP use to the partner and struggles to take PrEP discretely and consistently were identified from this work^30^, although these were not observed in the HPC due to most people being lost from the cascade at earlier steps.

Both young women and men reported highest motivation for, access to, and actual uptake of male condoms among all prevention methods analysed, reflecting the history of male condom programmes and availability within Zimbabwe and the wider region of southern Africa. Despite this, the gap between motivation and access was sizeable. A lack of self-efficacy – reporting lack of confidence to use male condoms regularly or to use them despite partner, peer, or family disapproval – was indicated as a barrier by the HPC, and has been found to be associated with lower odds of condom use in both men and women^21^. There was a large drop-off between the motivation and access to male condoms for both men and women, highlighting that access-related barriers need to be addressed.

VMMC levels reported are well below the target of 80% of 15-29 year-old men set out in Zimbabwe’s National HIV and AIDS Strategic Plan, with only 23% of men reporting full medical male circumcision^27^. Perceived negative consequences of VMMC were reported by 68% of men unmotivated to have VMMC. Lack of affordability – likely relating to time off work for the procedure and recovery – was the largest barrier to accessing VMMC although lack of easy access and lack of acceptable provision were also commonly reported with more than half of those with motivation but not access reporting a lack of easy access to VMMC services.

Lack of self-perceived future risk of HIV was a common barrier across prevention methods and has been shown to be associated with high HIV incidence^31^. Lack of accurate risk perception has been noted in young people in our study population in east Zimbabwe^31^. A lack of social acceptability was a commonly reported barrier across multiple prevention methods (male condoms, female condoms, VMMC) by both young men and women. Partner resistance was a commonly reported barrier by females for use of both female and male condoms.

Use of self-reported data could cause underestimation of the priority population (despite use of secret voting methods^22^) and overestimation of effective use of prevention methods. These analyses only assess a cross-section of risk behaviour and prevention method use, relying on the assumption that these remain the same in the future - a particular issue for young people whose sexual behaviour can change over short periods of time^32^. Analyses do not explore differences in the HPCs according to the type of self-reported risk behaviour. Only the main bars of the HIV-CPC were presented due to the complexity of populating explanatory barriers for multiple prevention methods in one cascade.

Despite these limitations, our findings demonstrate the value and utility of measuring the Harare HIV-CPC framework in a population survey. For AGYW and ABYM, they highlight the need for interventions in all parts of the cascades. Furthermore, the variations found between the gaps and barriers for different HIV prevention methods and priority populations point to a need for interventions which are specific to particular methods and populations as well as the local context. Nevertheless, some barriers are common across prevention methods and could be targeted by broader, cross cutting interventions. For example, appropriate community-level interventions could improve knowledge of risky sexual behaviours and reduce social unacceptability by addressing the stigma attached to use of prevention methods. Interventions which increase the privacy and confidentiality of prevention method providers, and public confidence in this privacy, could reduce unacceptable provision as a barrier. The role of partner as a barrier in the capacity to use prevention methods was consistently reported across multiple methods. Interventions to improve AGYW’s capacity to negotiate prevention method use with a partner and acceptance of use within a partnership could increase prevention method use.

## Conclusions

The HIV-CPC has enabled identification of barriers to motivation (particularly knowledge of PrEP, social acceptability of condoms and VMMC, and current and future risk perception), access (particularly availability of PrEP, and acceptable provision of all primary prevention methods), and, ultimately, effective use of primary HIV prevention methods (particularly the practical and social skills required to negotiate use of primary prevention methods with a sexual partner). These barriers vary by priority population and prevention method and could be targeted by interventions to improve effective use, including increasing motivation to use prevention methods, and removing fear of stigma and judgement of prevention method use in young men and women. High proportions of individuals engaging in risky sexual behaviour remain, indicating a need to improve HIV prevention method use to prevent acquisition of HIV. Even when young people are motivated and have access to prevention, barriers remain including lack of social skills and self-efficacy to negotiate prevention method use.

## Author Contributions

SG, TBH, CN, JWI-E and LM conceptualised the study. CN, BT, RM, and PM-M had major roles in collection and management of data used in this study. LM analysed the data. LM, SG and JWI-E contributed to interpretation of results. LM wrote the initial report which was reviewed and revised by all co-authors. All authors had final responsibility for the decision to submit for publication.

## Acknowledgements

We are grateful to the research team working at the Manicaland Centre for Public Health, as well as to the study participants in Manicaland, for their contribution to this study.

This work was supported by the Bill and Melinda Gates Foundation (BMGF) (INV-09999) and the National Institute of Mental Health (NIMH) (R01MH114562–01). LM, SG, TBH, JWI-E and CN acknowledge funding from the MRC Centre for Global Infectious Disease Analysis (reference MR/X020258/1), funded by the UK Medical Research Council (MRC). This UK funded award is carried out in the frame of the Global Health EDCTP3 Joint Undertaking.

## Data access statement

Due to the sensitive nature of data collected, including information on HIV status, treatment and sexual risk behaviour, the Manicaland Centre for Public Health does not make full analysis datasets publicly available. Summary datasets of household and background sociodemographic individual questionnaire data, covering rounds 1-8 (1998-2021) of the Manicaland General Population HIV Sero-Survey, are publicly available for download via the Manicaland Centre for Public Health website here - http://www.manicalandhivproject.org/data-access.html. Quantitative data used for analyses produced by the Manicaland Centre for Public Health are available on request following completion of a data access request form here - http://www.manicalandhivproject.org/data-access.html. Additionally, summary HIV incidence and mortality data spanning rounds 1-6 (1998-2013), created in collaboration with the ALPHA Network are available via the DataFirst Repository here - https://www.datafirst.uct.ac.za/dataportal/index.php/catalog/ALPHA/about

